# Reproduction number of monkeypox in the early stage of the 2022 multi-country outbreak

**DOI:** 10.1101/2022.07.26.22278042

**Authors:** Zhanwei Du, Zengyang Shao, Yuan Bai, Lin Wang, Jose L Herrera-Diestra, Spencer J. Fox, Zeynep Ertem, Eric H. Y. Lau, Benjamin J. Cowling

## Abstract

Monkeypox, a fast-spreading viral zoonosis outside of Africa in May 2022, has scientists on alert. We estimated the reproduction number to be 1.29 (95% CrI: 1.26, 1.33) by aggregating all cases in 70 countries as of July 22, 2022.

## Main Text

Monkeypox is a sylvatic zoonosis, transmitted by close contact and droplet exposure via droplets [1] and recognized as the most important orthopoxvirus infection after the eradication of smallpox [2]. It was first detected in monkeys in 1958 during an outbreak in an animal facility in Denmark, and the first human case was recorded in Central Africa in 1970 [3,4]. In the past decades, monkeypox usually occurred sporadically in forested parts of Central and West Africa, with two distinct genetic clades; the West African clade and Congo Basin clade [1], with cases only rarely detected outside of Africa. On May 7, 2022, the World Health Organization was informed of a confirmed case of monkeypox in the United Kingdom in an individual who traveled from the United Kingdom to Nigeria [1]. Eleven days later, Portugal, Spain, and the United States, where outbreaks have rarely occurred, reported 14, 18 and 1 cases of monkeypox, respectively. The monkeypox virus might have been spreading cryptically in these and other countries before the WHO declaration. By July 22, 2022, over 70 countries had reported a total of 16,313 cases of the monkeypox virus [5]. At that point, a rapid global response was then coordinated, including ring vaccination, following the atypical multi-country outbreak. The World Health Organization did not consider monkeypox to be a public health emergency of international concern after a meeting on June 23, 2022 [6], but reassessed the situation again and declared an emergency on July 23, 2022 [7].

The effective reproduction number (*R*_*e*_) and the incubation period are key epidemiological metrics to adjusting the response against the outbreaks caused by pathogens. They are defined as the expected number of secondary infections per primary infection in a partly susceptible population, and the duration between exposure and symptom onset of a case, respectively. In this study, we performed a systematic review to synthesize the evidence from published estimates of the effective reproduction number and incubation period for the monkeypox virus. We further estimated the effective reproduction number in the ongoing outbreak in six countries (including five countries with the confirmed cases exceeding 1000 and Portugal) between May and July 2022.

## Methods

### Search strategy and selection criteria

All searches were carried out on July 24, 2022 in PubMed, Embase, and Web of Science for articles published until July 24, 2022. Our search terms for incubation period and reproduction numbers of monkeypox include (#1) “monkeypox” OR “monkey pox” OR “MPXV”; (#2) “reproduct*” OR “incubation period” OR contact rate OR “transmissibility”. Our final search term was #1 AND #2. After reading the abstract and full text, we included the 11 studies that provided information about the incubation period and reproduction numbers of monkeypox. All data were extracted independently and entered in a standardized form by two co-authors (Z. S and Z. D). Conflicts over inclusion of the studies and retrieving the estimates of these variables were resolved by the author (Z. D.). Information was extracted on the estimates of the reproduction number of monkeypox. Other information such as study’s location and period were also extracted for each selected study.

### Reproduction number estimation analysis

Let denote the raw confirmed cases, collected from HealthMap [12]. To reduce the data noise, we adjust to be using the 7-day moving averages by

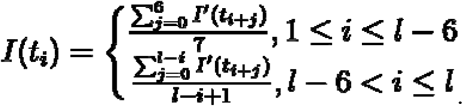

Following the subexponential model [13], the daily confirmed case is given by where and are the growth rate and the decelerating rate, respectively. The real-time reproduction number, *R*_*t*_, at time *t* is given by [13]

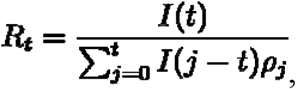

where is the discrete probability distribution of the generation interval. We assume the generation interval following the gamma distribution, with the mean value of 9.8 days and the standard deviation of 4 days [14]. We denote the effective reproduction number, *R*_*e*_, as *R*_*t*_ on the last study day.

Informed by the daily confirmed cases in the study region, we estimate the normal distributions for and, respectively, to minimize the residual sum of squares between the model output and the adjusted observed □(□) using the Levenberg–Marquardt algorithm [15]. We sampled 1000 pairs of □and □from their normal distributions independently, which are used to simulate □(□) and estimate the median and 95% CrI of *R*_*e*_ accordingly.

## Results

We identified 276 studies in total by searching PubMed, Embase, and Web of Science for articles published until 24 July 2022, in which 172 irrelevant studies were removed and excluded through title and abstract screening (**Figure 1**). A total of 11 studies were finally included in this review, which provides six *R*_*e*_ estimates in five regions (Congo, Central Africa, England, Portugal, and Spain) and seven estimates of incubation periods in five regions (Congo, Central and West Africa, the United States, Netherlands, and Germany). Two distinct genetic clades of the monkeypox virus were reported: the west African clade and the central African (Congo Basin) clade. Before 2022, *R*_*e*_ is estimated to be 0.08 (95% CI: 0.02, 0.22) and in the range from 0.21 to 0.58, for the West African clade and the Congo Basin clade, respectively (**Table 1**). But *R*_*e*_ of the West African clade increases in the range from 1.4 to 1.8 in the 2022 outbreak. The incubation period ranges from 5 to 41 days and from 8 to 14 days for the West African clade and the Congo Basin clade, respectively (**Table 2**). We further estimate the effective reproduction number in outbreaks as of July 22, 2022 (**Figure 2, Table 3**). For those six countries with increasing confirmed cases, the United States has the highest *R*_*e*_ estimated as 1.55 (95% CrI: 1.42, 1.73). By aggregating all cases in 70 countries, *R*_*e*_ is estimated to be 1.29 (95% CrI: 1.26, 1.33), larger than four of the six countries with increasing confirmed cases. Compared to earlier *R*_*e*_ estimates (**Table 1, Table S2**), transmission may have slowed down recently with increased awareness of the monkeypox epidemic (**Table 3**), though with *R*_*e*_ >1 further spread is likely without more stringent containment measures. In addition, we conduct a sensitivity analysis of *R*_*e*_ by using unadjusted daily confirmed case data (**Table S1**) or adjusted cases before peaking incidence for four study countries (**Table S2**).

**Table 1.**
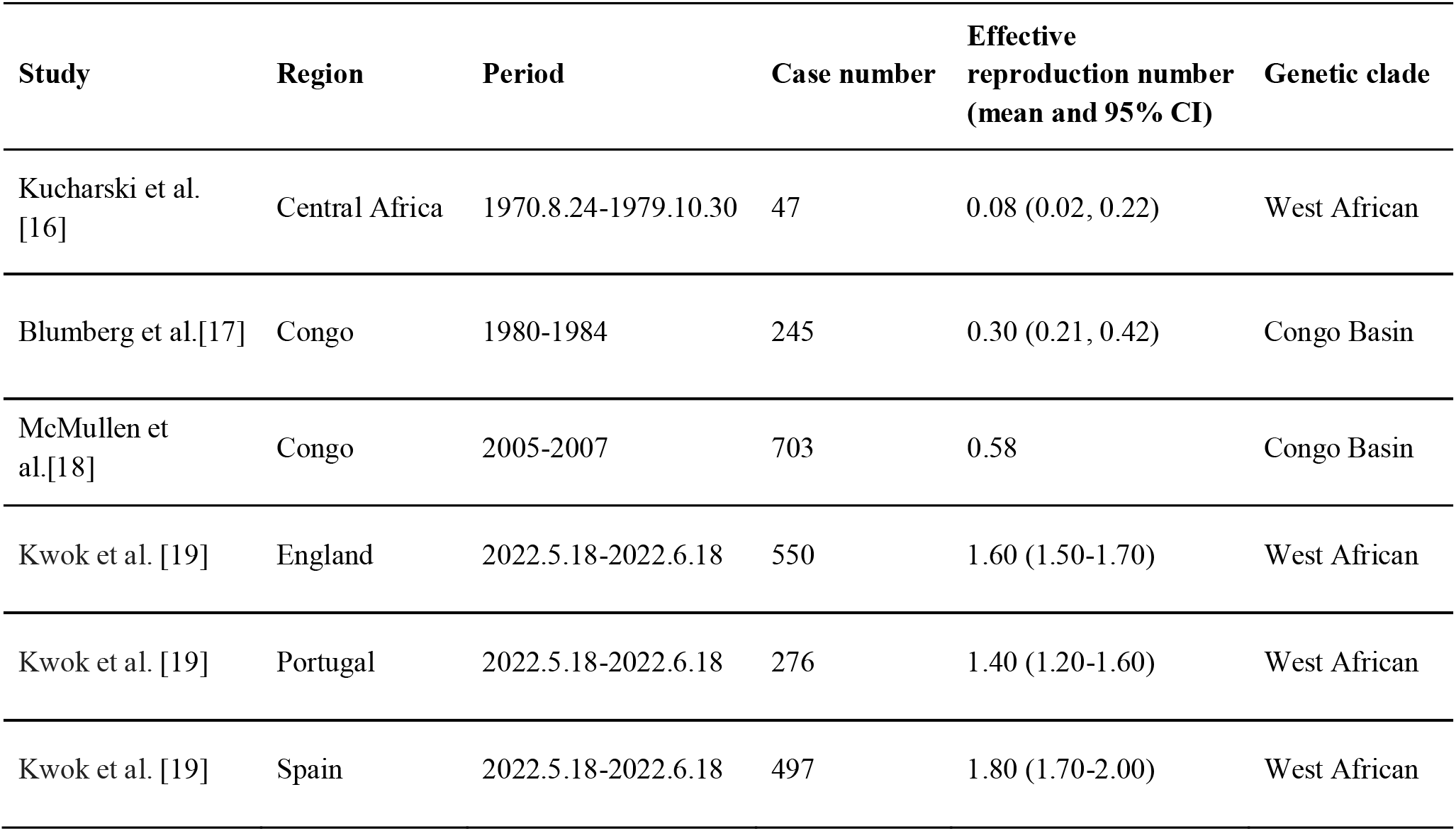
Summary of included studies for the effective reproduction number of monkeypox.

**Table 2.**
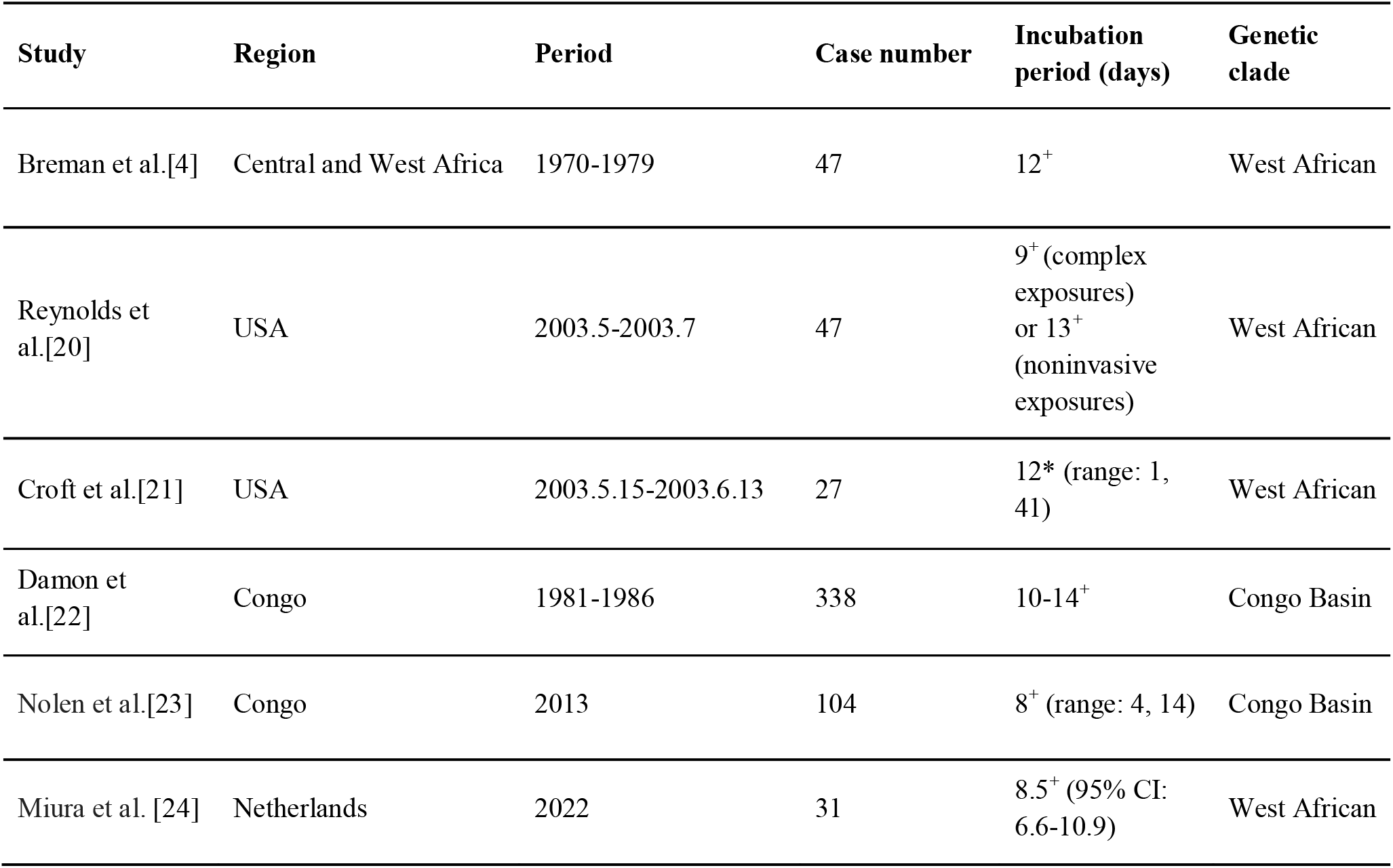
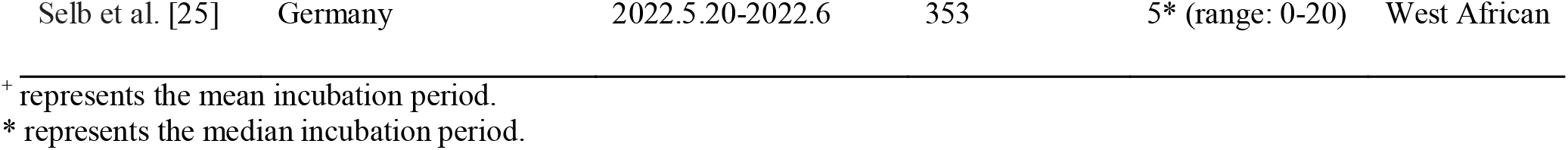
Summary of included studies for the incubation period of monkeypox.

**Table 3.** The estimated effective reproduction number of monkeypox informed by adjusted daily confirmed cases.

| $\hat{R}_t$<br>(median and 95% CrI) | Case number | Period | Region |
| --- | --- | --- | --- |
| 1.55<br>(1.42, 1.73) | 2581 | 2022.5.18-2022.7.22 | United States |
| 1.36<br>(1.24, 1.60) | 1562 | 2022.5.19-2022.7.22 | France |
| 1.20<br>(1.17, 1.23) | 2268 | 2022.5.19-2022.7.22 | Germany |
| 1.18<br>(1.10, 1.31) | 3125 | 2022.5.18-2022.7.22 | Spain |
| 1.13<br>(1.10, 1.16) | 2115 | 2022.5.6-2022.7.22 | England |
| 1.02<br>(1.00, 1.04) | 588 | 2022.5.17-2022.7.22 | Portugal |
| 1.29<br>(1.26, 1.33) | 16313 | 2022.5.6-2022.7.22 | All |

**Figure 1.**
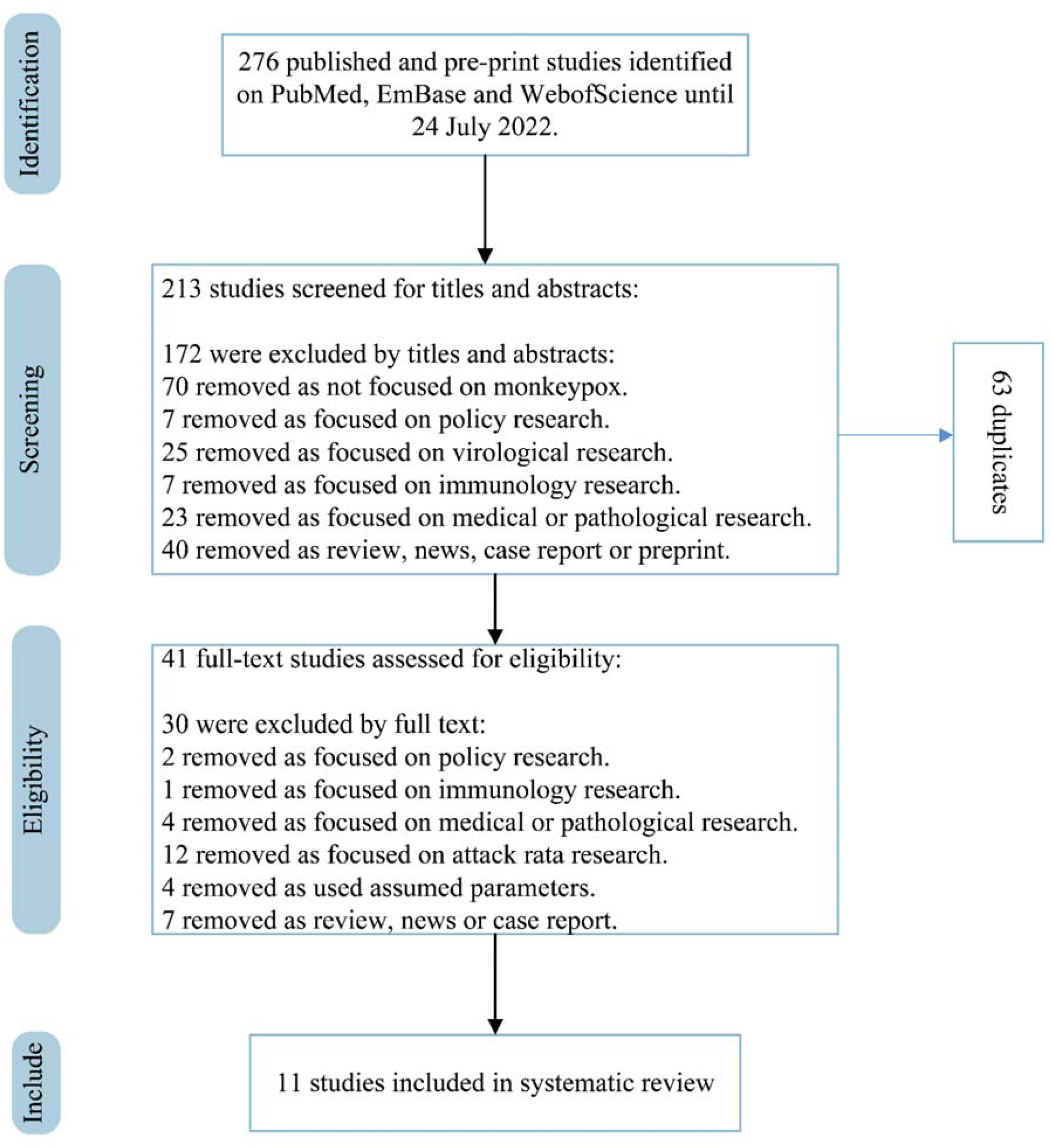
PRISMA (Preferred Reporting Items for Systematic Reviews and Meta-Analyses) flow diagram for the studies used to obtain studies that reported incubation period and reproduction numbers.

**Figure 2.**
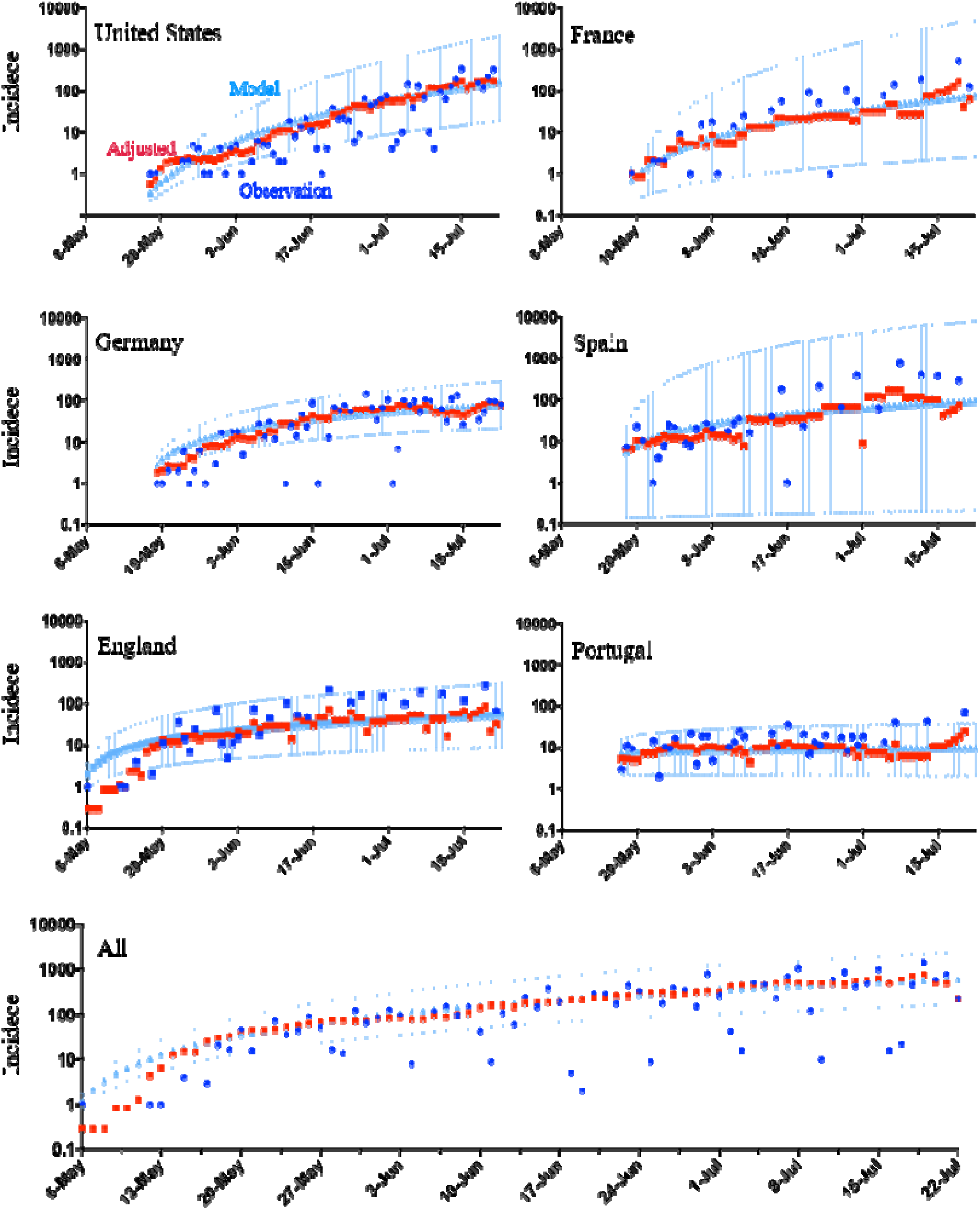
Estimated daily confirmed cases during the period between May 6 to July 22, 2022. We estimate the mean and 95% CI of daily confirmed cases, using the sub-exponential model informed by the adjusted (smoothed) observation from the actual observation across five countries and globally. The X-axis and Y-axis denote the reporting date and the daily cases, respectively.

## Discussion

In the 2022 global outbreak, the multi-country monkeypox virus from the West African clade may have found its global march outward into highly interconnected men who have sex with men (MSM) networks, where it can spread via unique behavioral pattern different from the general population [8,9]. For example, in the United Kingdom, there were 54 confirmed cases of monkeypox virus reported in London between May 14 and May 25, 2022, all of which occurred in MSM [10]. Male sexual encounters may play a role in transmission of monkeypox viruses [8], perhaps due to high viremia, local inflammation, imperfection of the blood-testis barrier, testis as an immunologically privileged site, and replication in accessory glands [11]. Increased propensity to seek healthcare among MSM may also contribute to increased diagnoses in this group to date.

While we believe that our qualitative findings are robust, our estimates have several limitations. We may overestimate the infection risks in the 2022 outbreaks. The modes of transmission in the reviewed studies are mainly in a general population, which is different from that in the ongoing outbreak in 2022. The estimated R_e_ has a value over one across countries; however, it would not denote a risk for the general population, but instead, it would reflect the risk faced by the MSM community.

In conclusion, multiple estimates of the reproduction number have been published for monkeypox. Reliable estimates of its reproduction numbers in an epidemic will aid accurate assessment of the impact of control efforts and the potential need to curtail the spread of monkeypox in both the MSM community and the general population.

## Data Availability

All data are collected from open source with detailed description in Section Method.

## Acknowledgments

We acknowledge the financial support from the AIR@InnoHK Programme from Innovation and Technology Commission of the Government of the Hong Kong Special Administrative Region, the Collaborative Research Fund (Project No. C7123-20G) of the Research Grants Council of the Hong Kong SAR Government, National Natural Science Foundation of China (grant no. 72104208), and Health and Medical Research Fund, Food and Health Bureau, Government of the Hong Kong Special Administrative Region (grant no. 21200632). The funders of the study had no role in study design, data collection, data analysis, data interpretation, or writing of the report. All code to perform the analyses and generate the figures in this study are available from the corresponding author upon reasonable request.

## Author Contributions

ZD, ZS, YB, LW, and BJC: conceived the study, designed statistical and modelling methods, conducted analyses, interpreted results, wrote and revised the manuscript; JLHD, SJF, and EHYL: interpreted results and revised the manuscript.

## Data availability

All data are collected from open source with detailed description in the Methods section.

## Code availability

Code used for data analysis is freely available upon request.

## Appendix

**Table S1.**
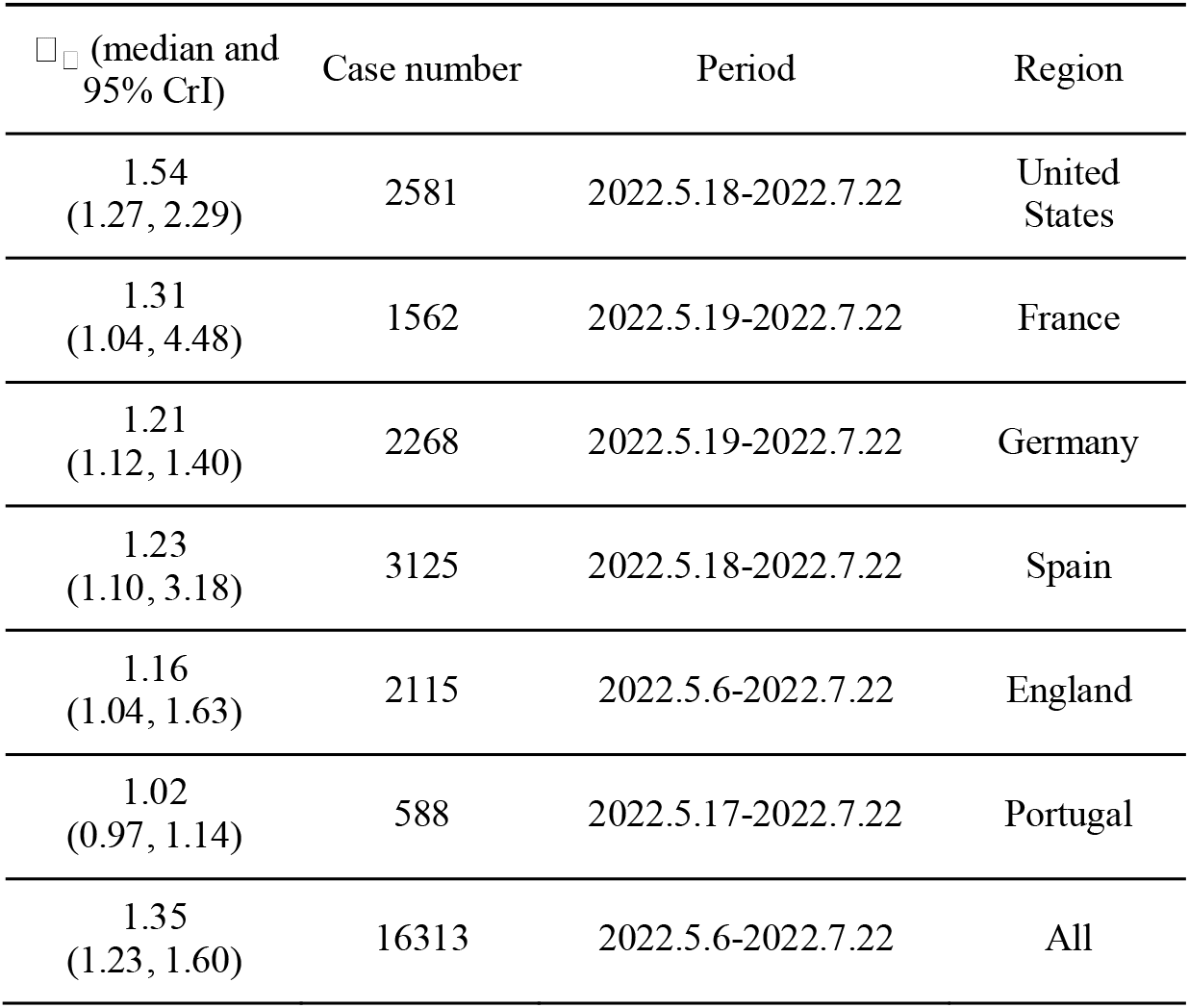
The estimated effective reproduction number of monkeypox using the unadjusted daily confirmed case data over regions.

**Table S2.** The estimated effective reproduction number of monkeypox using the adjusted daily confirmed case data before the peaking of incidences for each of four study countries. Given the stable trend of incidence in the four countries (Germany, Spain, England, and Portugal), we estimate the effective reproduction number before the peaking incidence.

| $\hat{R}_t$<br>(median and 95% CrI) | Case number | Period | Region |
| --- | --- | --- | --- |
| 1.96<br>(1.64, 2.55) | 763 | 2022.5.19-2022.6.27 | Germany |
| 2.48<br>(1.51, 5.65) | 736 | 2022.5.18-2022.6.23 | Spain |
| 1.81<br>(1.16, 4.56) | 766 | 2022.5.6-2022.6.20 | England |
| 1.29<br>(1.13, 1.60) | 276 | 2022.5.17-2022.6.17 | Portugal |

